## Supplementary information for "Characterizing trachoma elimination using serology"

### Supplementary Information Text

#### *Clinical assessment and serological testing*

Clinical examination was undertaken by trained field graders using the WHO simplified grading system [1]. Conjunctival swabs were passed over the conjunctiva three times, rotating 120° between passes, and tested for *C. trachomatis* DNA using PCR either as individual or pooled samples, as previously described [2]. Blood spots for serology assays were collected onto filter papers and most often tested in a multiplex bead assay for IgG antibodies to the Pgp3 and CT694 antigens, as previously described [3]. For this, antigen-coupled beads were mixed with control sera and blood spot eluates and total IgG determined by reading the beads on a Luminex instrument (Luminex Corp., Austin, TX) and converting the signal readout to median fluorescence intensity (MFI) with background levels subtracted out (MFI-BG). Seropositivity thresholds were generated using receiver-operating characteristic curves of specimens of previously classified positive or negative samples [4]. Other established platforms for detection of anti-Pgp3 antibody, ELISA and lateral flow assays (LFA), were also used in various contexts, as previously described [5–9]. The different serology testing platforms are generally closely comparable [4]. Thresholds to determine sero-positivity were defined for each study or dataset separately.

Further details of study design, clinical examination, sampling methodology and laboratory procedures can be found in publications for the respective individual studies (Table S1).

#### *Estimation of seroconversion rate within a generalized linear model using maximum likelihood*

The seroconversion rate (SCR) from a current status, single-rate catalytic model assuming no seroreversion is closely tied to the slope of age-seroprevalence curve. The SCR is equal to the slope of the age-seroprevalence curve divided by the complement of the seroprevalence at age A = a. It can be shown that the SCR based on this model can be estimated as the exponentiated intercept from a generalized linear model with binomial error structure and a complementary log-log link:

$$log[-\log(1-P(Y=1|A))] = log\lambda + logA$$

where Y represents individual-level serostatus (1: seropositive, 0: seronegative or equivocal), A is the child's age in years, and λ is the SCR. The SCR provides an estimate of the force of infection, an epidemiological parameter that denotes the rate at which susceptible individuals in the population become infected.

***Reversible catalytic model***

Next, we extended the model above to allow for seroreversion. Cross-sectional data do not contain sufficient information to reliably estimate both seroreversion and seroconversion rates. We fit a binomial maximum likelihood model for seroconversion with fixed a seroreversion rate of 6.6 per 100 person-years based on a longitudinal cohort in Kongwa, United Republic of Tanzania, monitored in the absence of MDA [10]:

$$P(Y = 1|A) = \frac{\lambda}{\lambda + \rho}[1 - e^{-(\lambda+\rho)(A)}]$$

where ρ is the assumed seroreversion rate.

As an internal validity check, we compared the SCR estimates from the simple and reversible models.

***Alternative categories based on elimination of transmission***

The primary analysis categorized EUs based on their need for population-level trachoma-specific interventions. In many cases, populations could reach a level of trachoma control that would not justify further large-scale interventions, such as antibiotic mass drug administration (MDA), yet still have underlying detectable ocular *C. trachomatis* transmission. We conducted an alternative analysis that categorized EUs by level of *C. trachomatis* transmission defined using clinical data (prevalence of trachomatous inflammation—follicular, TF), prevalence of infection by PCR, and programmatic context such as recent MDA application, with a goal of identifying a subgroup of EUs that had likely achieved interruption of transmission. A caveat of the analysis is that there are no formal benchmarks or international guidance for documenting elimination of ocular *C. trachomatis* transmission, but we report these results as an illustrative example for how the problem could be approached. It also provides an example of how the methodology used can generalize to include multiple transmission categories (not only two as in the primary analysis).

We created three categories: (i) `Probable interruption of transmission` – (n=14) – which comprised EUs with population-level TF of <5% in 1-9-year-olds, ocular infection prevalence of <1%, and no recent or planned MDA treatments for a minimum of one year prior to the survey, as MDA can transiently reduce infection prevalence. (ii) `Endemic` – (n=7) – EUs with unambiguously high transmission, as evident by high TF prevalence (>10%), high infection prevalence (>5%) and where MDA programs were in place or additional MDA treatments were planned. (iii) `Near interruption of transmission` – (n=14) – comprising EUs with intermediate clinical and PCR measures that were not at either extreme described above. The near interruption of transmission EUs were thought as being on the path to interruption of transmission but with unusual epidemiology (e.g., higher prevalence of clinical signs or infection than would be expected) or requiring an additional round of MDA or monitoring to assess disease status. Finally, EUs with IgG measurements that lacked clinical or PCR measures were used as an independent, held-out (or test-set) sample to assess the methodology (n=13). It should be noted that the infection thresholds used here to demarcate the interruption of transmission and endemic categories are strictly hypothetical and should not be interpreted as factual, or as representing the views, decisions or policies of any organization contributing to the fight for global trachoma elimination.

We used the same approach as the primary analysis to make probability statements for each transmission category, a 3-component mixture model for the pooled distributions of SCR estimates. This approach assumed that each MCMC estimate is drawn independently from a multinomial distribution made of three components parallel to the three transmission categories defined above, $k \in \{1, 2, 3\}$. So, for each transmission category ($C_k$: probable interruption of transmission, near interruption of transmission, endemic) and SCR estimate, $x \epsilon \mathbb{R}$, we compute the posterior probability, $p(C_k|x)$, using the Bayes rule:

$$p(C_k|x) \propto p(C_k) * \frac{p(x|C_k)}{p(x)}$$

where $p(C_k)$ is the prior probability that a population is in transmission category $C_k$; and $p(x|C)$, is the likelihood evaluated as empirical probability density function at each MCMC draw $x$. $p(x)$ denotes the marginal likelihood or normalizing constant for the posterior density obtained by integrating the products of the likelihood, $p(x|C_k)$, and the prior probability. That is, the sum of the products of the density function and prior probability for each $k$,

$$\sum_{k=1}^{3} \omega_k f_k(x|C_k)$$

$p(x|C)/p(x)$ forms the likelihood ratio in the Bayesian mixture model. The prior probabilities were defined such that they sum up to one, i.e., $\sum_k \omega_k = 1$. We used a general quadratic function $f(x) = ax^2 + bx + c$ such as,

$$a = b = p\left(C_{k=Probable\ interruption\ of\ transmission}\right)$$

$$c = p\left(C_{k=Probable\ interruption\ of\ transmission}\right) - 1$$

The positive root, $x$, of the quadratic equation above was used to determine the prior probabilities of the near interruption of transmission and endemic categories as,

$$p\left(C_{k=Near\ interruption\ of\ transmission}\right) = a * x$$

$$p(C_{k=Endemic}) = a * x^2$$

Of the 48 EUs, 35 had information on TF and infection prevalence and could be classified into the three transmission categories defined above. The remaining unclassified EUs in this supplementary analysis (n=13) were treated as a held-out set of surveys and used with the mixture model approach to estimate their posterior probability of each transmission category.

SCR estimates were generally well separated between trachoma transmission categories defined using clinical and infection measures. SCRs were low among the EUs in the probable interruption of transmission category, with the median SCR in the age group 1-5 years estimated between 0.1 (95% CrI, 0.0 to 0.2) and 0.9 (0.5 to 1.6) per 100 person-years. In the near interruption of transmission category, the estimated SCRs ranged from 0.5 (0.2-1) to 4.1 (3.1-5.5) per 100 person-years. There was substantial heterogeneity in SCR estimates among EUs in the endemic transmission category, with median estimates as high as 17.6 (13.6-22.3) per 100 person-years.

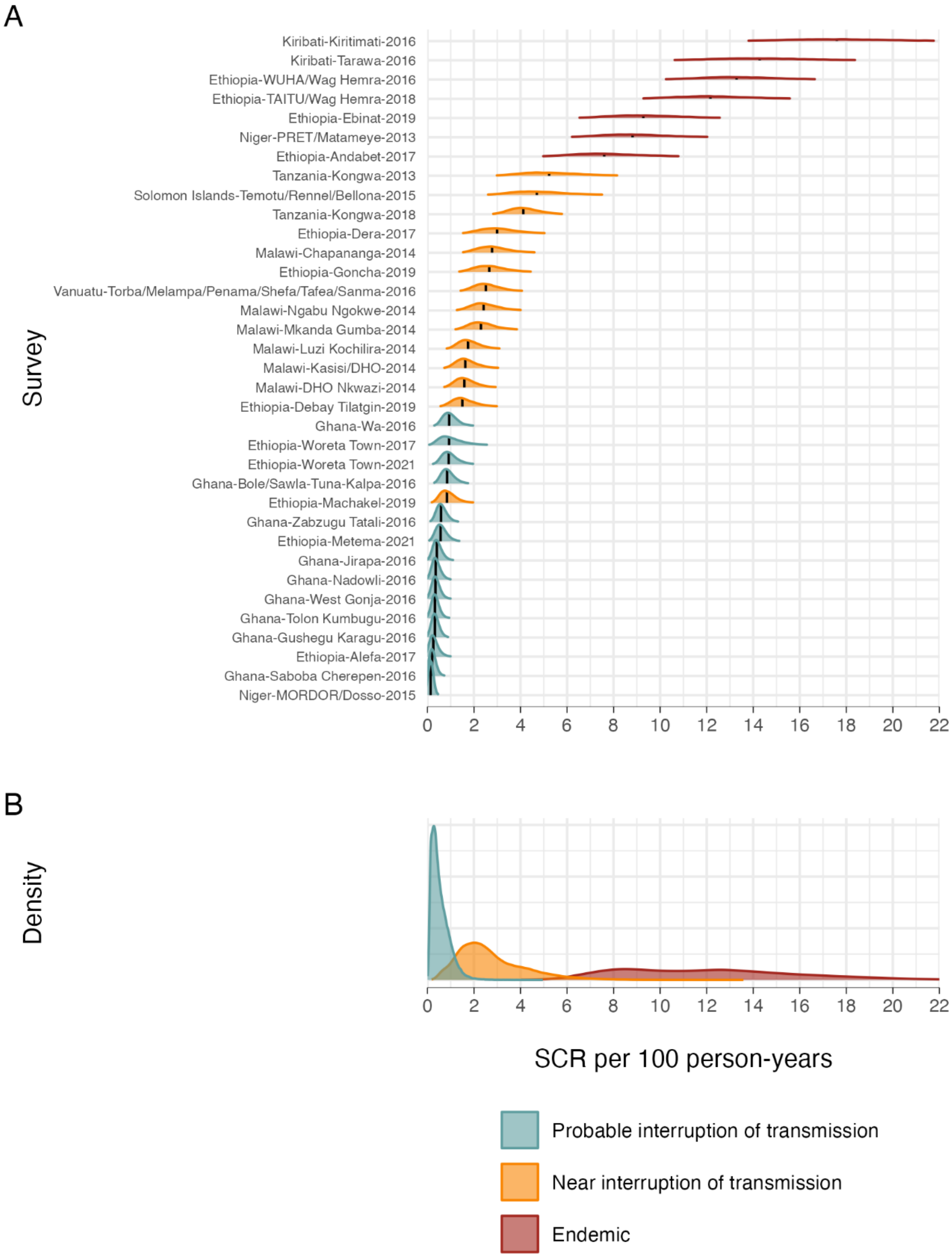
A
Survey
Kiribati-Kiritimati-2016
Kiribati-Tarawa-2016
Ethiopia-WUHA/Wag Hemra-2016
Ethiopia-TAITU/Wag Hemra-2018
Ethiopia-Ebinat-2019
Niger-PRET/Matameye-2013
Ethiopia-Andabet-2017
Tanzania-Kongwa-2013
Solomon Islands-Temotu/Rennel/Bellona-2015
Tanzania-Kongwa-2018
Ethiopia-Dera-2017
Malawi-Chapananga-2014
Ethiopia-Goncha-2019
Vanuatu-Torba/Melampa/Penama/Shefa/Tafea/Sanma-2016
Malawi-Ngabu Ngokwe-2014
Malawi-Mkanda Gumba-2014
Malawi-Luzi Kochilira-2014
Malawi-Kasisi/DHO-2014
Malawi-DHO Nkwazi-2014
Ethiopia-Debay Tilatgin-2019
Ghana-Wa-2016
Ethiopia-Woreta Town-2017
Ethiopia-Woreta Town-2021
Ghana-Bole/Sawla-Tuna-Kalpa-2016
Ethiopia-Machakel-2019
Ghana-Zabzugu Tatali-2016
Ethiopia-Metema-2021
Ghana-Jirapa-2016
Ghana-Nadowli-2016
Ghana-West Gonja-2016
Ghana-Tolon Kumbugu-2016
Ghana-Gushegu Karagu-2016
Ethiopia-Alefa-2017
Ghana-Saboba Cherepen-2016
Niger-MORDOR/Dosso-2015
0 2 4 6 8 10 12 14 16 18 20 22
B
Density
0 2 4 6 8 10 12 14 16 18 20 22
SCR per 100 person-years
Probable interruption of transmission
Near interruption of transmission
Endemic

With a moderately informative prior of 80% probability of elimination, the posterior probability that an EU is in the post elimination category exceeds 90% when the SCR is = 1.0 per 100 person-years.

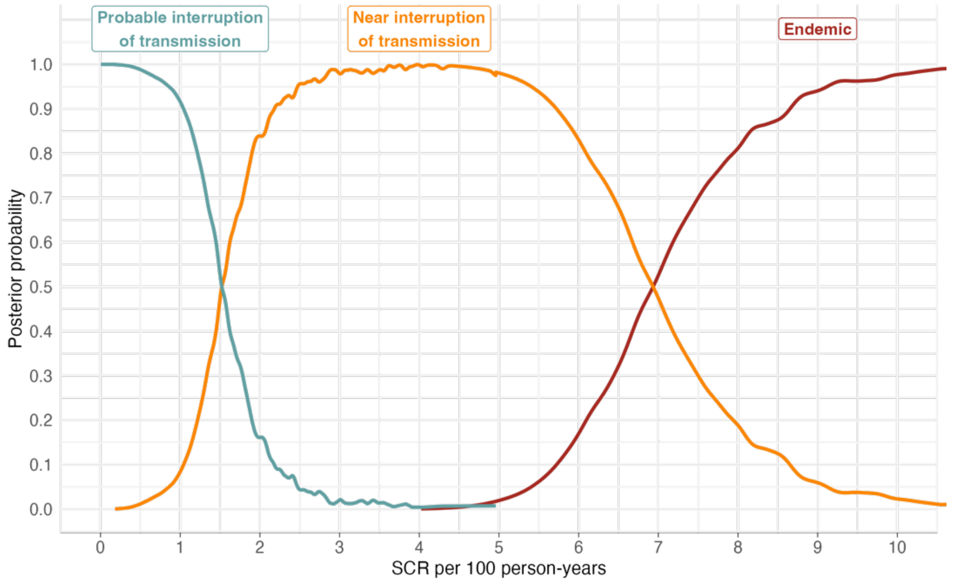

For each held-out EU, we calculated the posterior probability of its membership in each transmission category given its SCR distribution, assuming informative prior probabilities of 80% elimination, 17% near elimination, and 3% endemic categories.

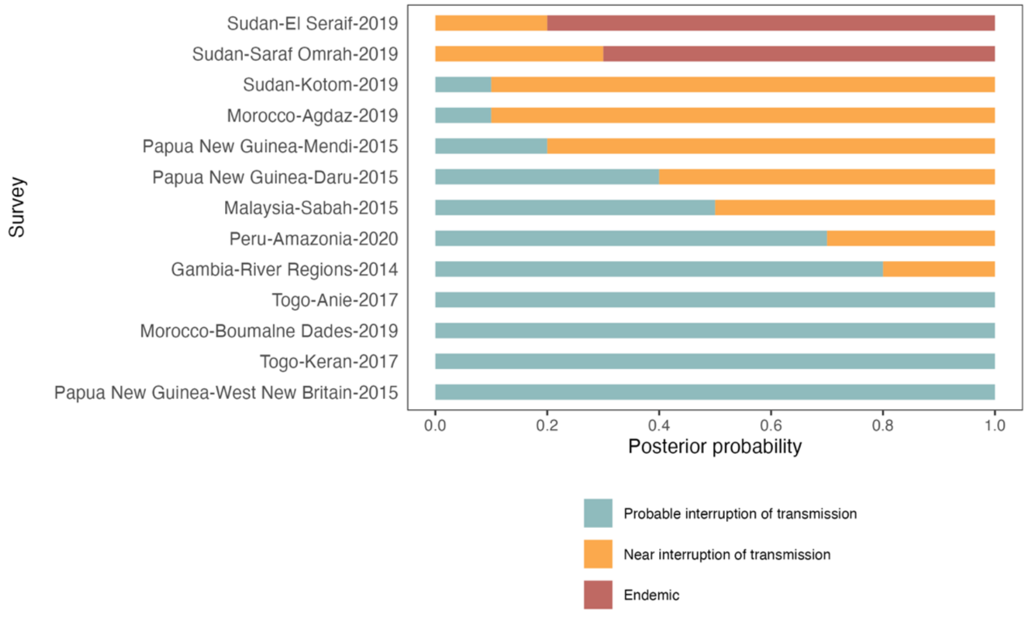

### Supplementary Information Tables

**Table S1**. Characteristics of the studies and sources of datasets analyzed here.

| Country | Region / District / Evaluation unit(s) | Year of survey | Serology assay | Relevant publication(s) |
|---|---|---|---|---|
| Ethiopia | Alefa | 2017 | MBA | PMID: 33200728 |
| Ethiopia | Andabet | 2017 | MBA | PMID: 33200728 |
| Ethiopia | Debay Tilatgin | 2019 | MBA | In peer review |
| Ethiopia | Dera | 2017 | MBA | PMID: 33200728 |
| Ethiopia | Ebinat | 2019 | MBA | In peer review |
| Ethiopia | Goncha | 2019 | MBA | In peer review |
| Ethiopia | Machakel | 2019 | MBA | In peer review |
| Ethiopia | Metema | 2021 | MBA | PMID: 38386689 |
| Ethiopia | Wag Hemra | 2018 | MBA | PMID: 32877398 |
| Ethiopia | Wag Hemra | 2016 | MBA | PMID: 34919861 |
| Ethiopia | Woreta Town | 2017 | MBA | PMID: 33200728 |
| Ethiopia | Woreta Town | 2021 | MBA | PMID: 38386689 |
| The Gambia | Lower & Upper River Regions | 2014 | ELISA | PMID: 28099433 |
| Ghana | Bole; Sawla-Tuna-Kalpa | 2016 | MBA | PMID: 30550537 |
| Ghana | Gushegu Karagu | 2016 | MBA | PMID: 30550537 |
| Ghana | Jirapa | 2016 | MBA | PMID: 30550537 |
| Ghana | Nadowli | 2016 | MBA | PMID: 30550537 |
| Ghana | Saboba Cherepen | 2016 | MBA | PMID: 30550537 |
| Ghana | Tolon Kumbugu | 2016 | MBA | PMID: 30550537 |
| Ghana | Wa | 2016 | MBA | PMID: 30550537 |
| Ghana | West Gonja | 2016 | MBA | PMID: 30550537 |
| Ghana | Zabzugu Tatali | 2016 | MBA | PMID: 30550537 |
| Kiribati | Kiritimati | 2016 | MBA | PMID: 28898240 |
| Kiribati | Tarawa | 2016 | MBA | PMID: 28898240 |
| Malawi | Chapananga | 2014 | ELISA | PMID: 31658258 |
| Malawi | DHO Nkwazi | 2014 | ELISA | PMID: 31658258 |
| Malawi | Kasisi/DHO | 2014 | ELISA | PMID: 31658258 |
| Malawi | Luzi Kochilira | 2014 | ELISA | PMID: 31658258 |
| Malawi | Mkanda Gumba | 2014 | ELISA | PMID: 31658258 |
| Malawi | Ngabu Ngokwe | 2014 | ELISA | PMID: 31658258 |
| Malaysia | Sabah | 2015 | MBA | PMID: 36388287 |
| Morocco | Agdaz | 2019 | MBA | PMID: 35344929 |
| Morocco | Boumalne Dades | 2019 | MBA | PMID: 35344929 |

| | | | | |
|---|---|---|---|---|
| Niger | Dosso | 2015 | MBA | PMID: 31237871 |
| Niger | Matameye | 2013 | MBA | PMID: 27956455 |
| Papua New Guinea | Daru | 2015 | MBA | PMID: 31965155 |
| Papua New Guinea | Mendi | 2015 | MBA | PMID: 31965155 |
| Papua New Guinea | West New Britain | 2015 | MBA | PMID: 31965155 |
| Peru | Amazonia region | 2020 | MBA | In peer review |
| Solomon Islands | Temotu; Rennel; Bellona | 2015 | MBA | PMID: 29588922 |
| Sudan | El Seraif | 2019 | MBA | PMID: 38507810 |
| Sudan | Kotom | 2019 | MBA | PMID: 38507810 |
| Sudan | Saraf Omrah | 2019 | MBA | PMID: 38507810 |
| Tanzania | Kongwa | 2013 | MBA | PMID: 30543311 |
| Tanzania | Kongwa | 2018 | MBA | PMID: 33861754 |
| Togo | Anie | 2017 | MBA | PMID: 33790370 |
| Togo | Keran | 2017 | MBA | PMID: 33790370 |
| Vanuatu | Torba; Malampa; Penama; Shefa; Tafea; Sanma | 2016 | MBA | PMID: 32017971 |

* PMID = PubMed ID. ELISA = enzyme-linked immunosorbent assay. MBA = multiplex bead assay.

**Table S2**. Evaluation unit-level seroconversion rates (SCR) per 100 person-years among children 1–5-years-old corresponding with different trachoma programmatic decisions (Action not needed, Action needed) across varying prior probability assumptions and levels of certainty based on the posterior probability of each category.

| **Prior Probability** | **Level of Certainty** | **SCR threshold Action not needed** | **SCR threshold Action needed** |
|---|---|---|---|
| 80% Action not needed / 20% Action needed | 99% | 1.3 | 7.4 |
| | 95% | 1.8 | 5.1 |
| | 90% | 2.2 | 4.5 |
| | 80% | 2.8 | 4.1 |
| 50% Action not needed / 50% Action needed | 99% | 1.0 | 7.4 |
| | 95% | 1.4 | 4.2 |
| | 90% | 1.6 | 3.8 |
| | 80% | 1.9 | 3.4 |
| 20% Action not needed / 80% Action needed | 99% | * | 7.4 |
| | 95% | 1.0 | 3.6 |
| | 90% | 1.2 | 3.2 |
| | 80% | 1.4 | 2.8 |

* There was no SCR threshold value for the `Action not needed` programmatic decision corresponding with 99% certainty under a weak prior probability of 20% Action not needed.

### Supplementary Information Figures

**Fig S1. Seroconversion rate (SCR) per 100 person-years in 1–5-year-olds (N=41,168) – using a catalytic model assuming constant force of infection**. Density distributions of the SCR for 48 evaluation units (EUs). The black vertical line shows the median estimate, and the density distributions depict the uncertainty about the median. EUs are colored by categories based on programmatic responses (Methods) and ordered by increasing median SCR value.

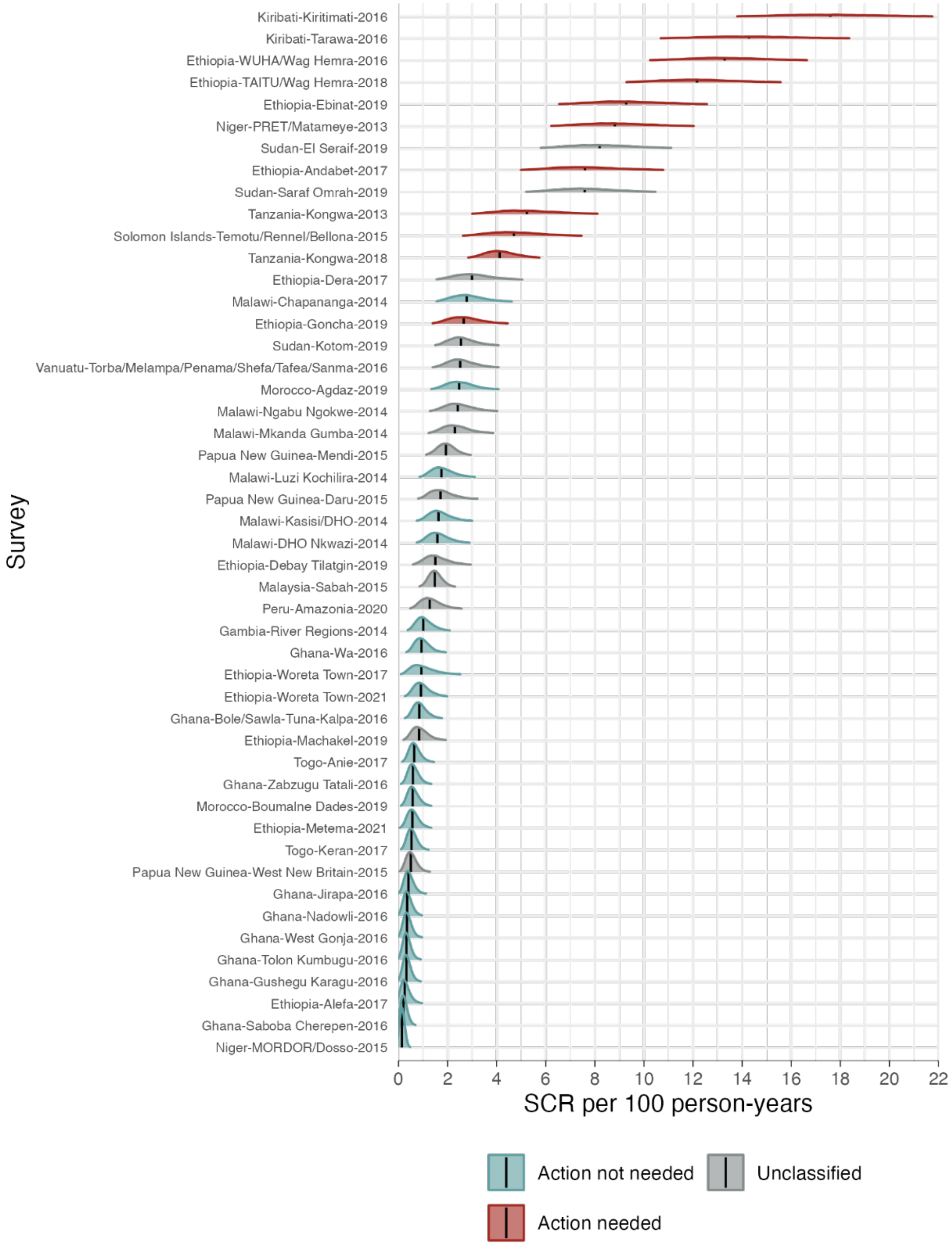

**FigS2. Relationship between the seroconversion rate (SCR) and seroprevalence at the evaluation unit (EU) level among 1–5-year-olds**. EUs are colored by transmission categories used in the main analysis (N=48 EUs). The right panel provides a zoomed view of estimates for populations approaching elimination, illustrating a highly linear relationship between serological summary measures. A linear regression fit to the 41 EUs with seroprevalence ≤25% led to coefficients $SCR \sim 0 + 0.363 \times Seroprev$ ($R^2$ =0.99), with SCR in units per 100 and seroprevalence in percentage points.

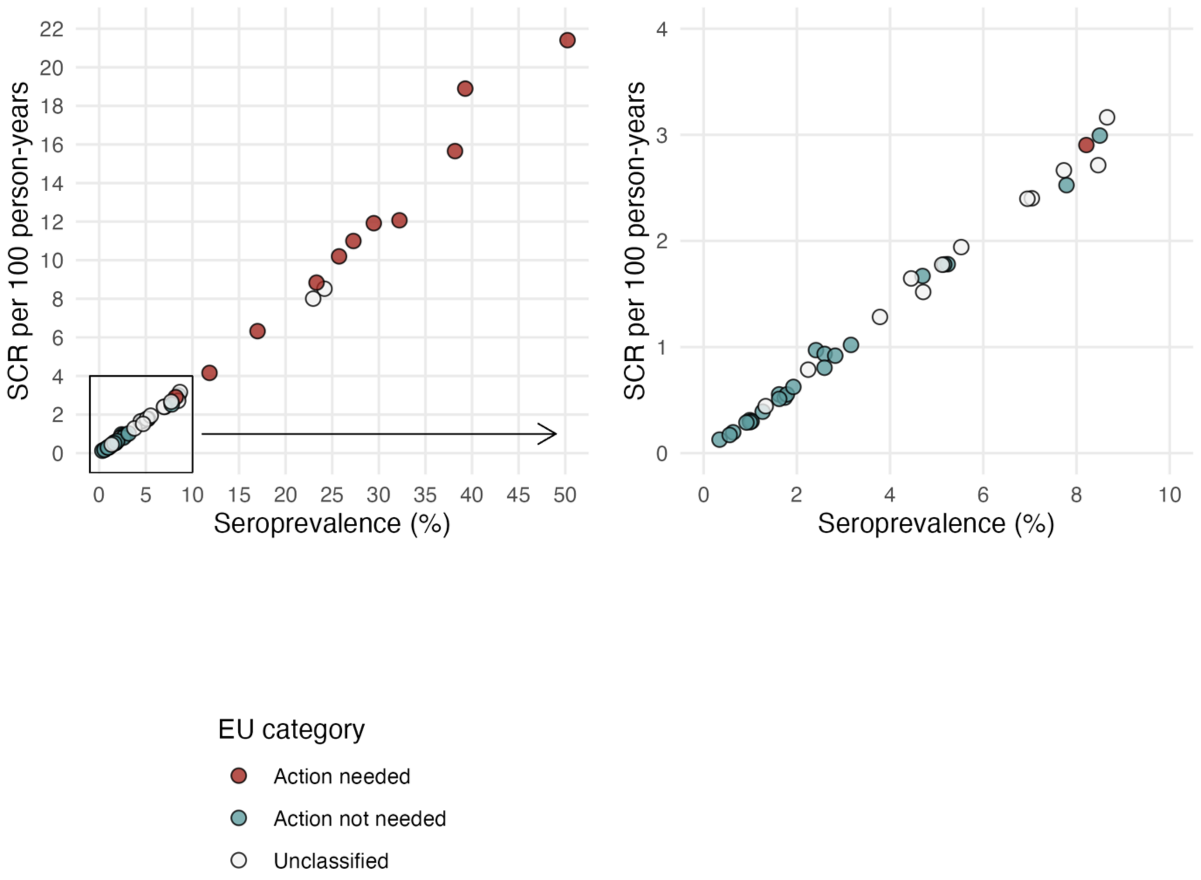

**Fig S3. Seroprevalence at evaluation unit (EU) level in 1–5-year-olds.** Seroprevalence was estimated by fitting generalized linear mixed models using Bayesian Markov chain Monte Carlo methods (see Methods). **(A)** Posterior density distributions of seroprevalence for each EU. The median estimate is showed by a black vertical line and the density distributions depict the uncertainty about the median. EUs are ordered by increasing median value and colored by programmatic action categories of whether additional trachoma-specific interventions are needed (Methods). **(B)** Pooled density distributions for each category.

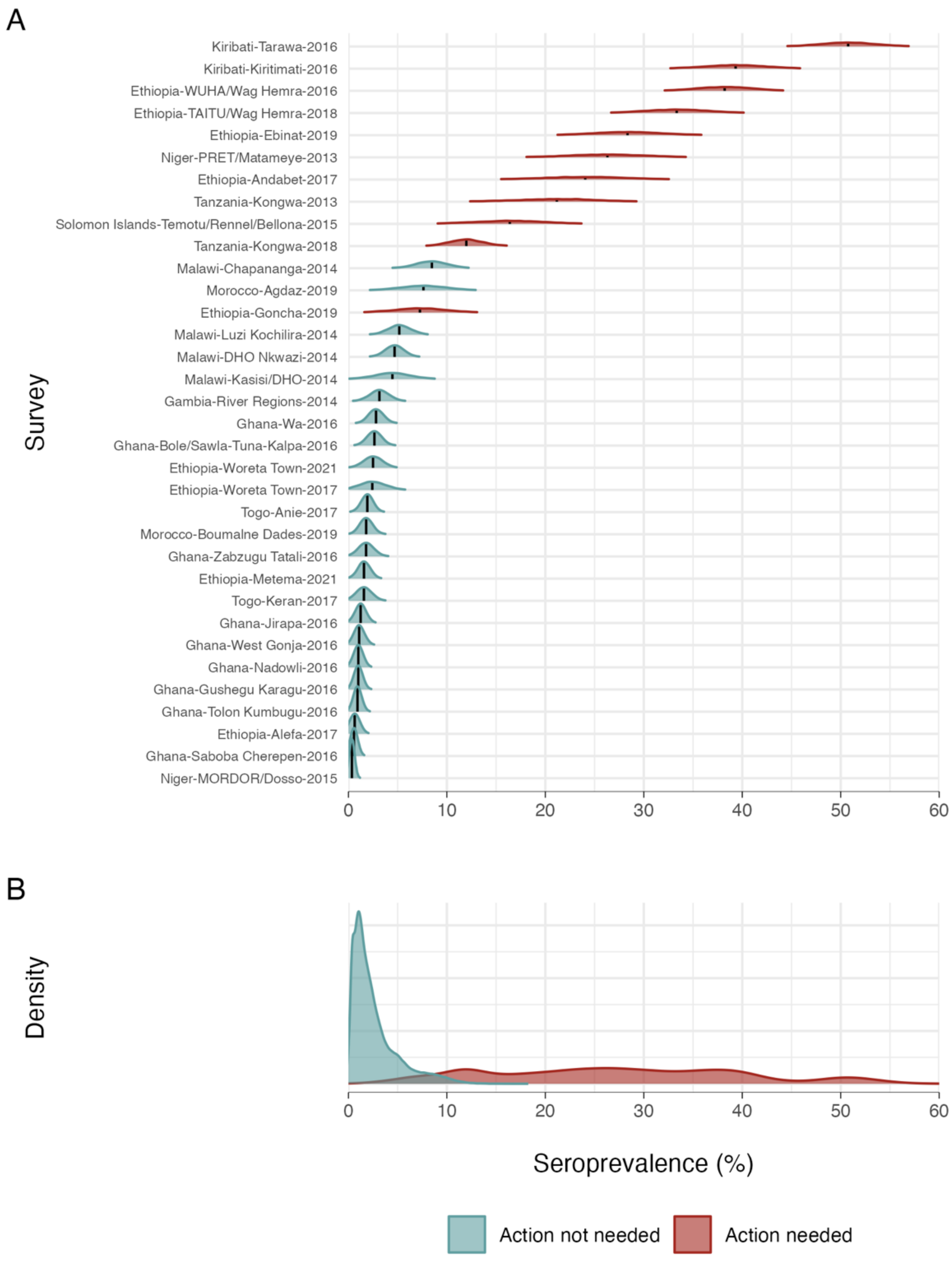

**Fig S4. Sensitivity analysis of posterior probability of categories given the seroconversion rate.** Evaluation units were grouped into Action not needed and Action needed categories and the pooled SCR estimates used in a Bayesian mixture model to obtain posterior probabilities (Methods). The plots depict posterior probabilities of each category for a range of weakly and moderately informative prior probabilities. We considered stronger priors for the Action not needed category in panel **(A)**, and stronger priors for the Action needed category in panel **(B)**.

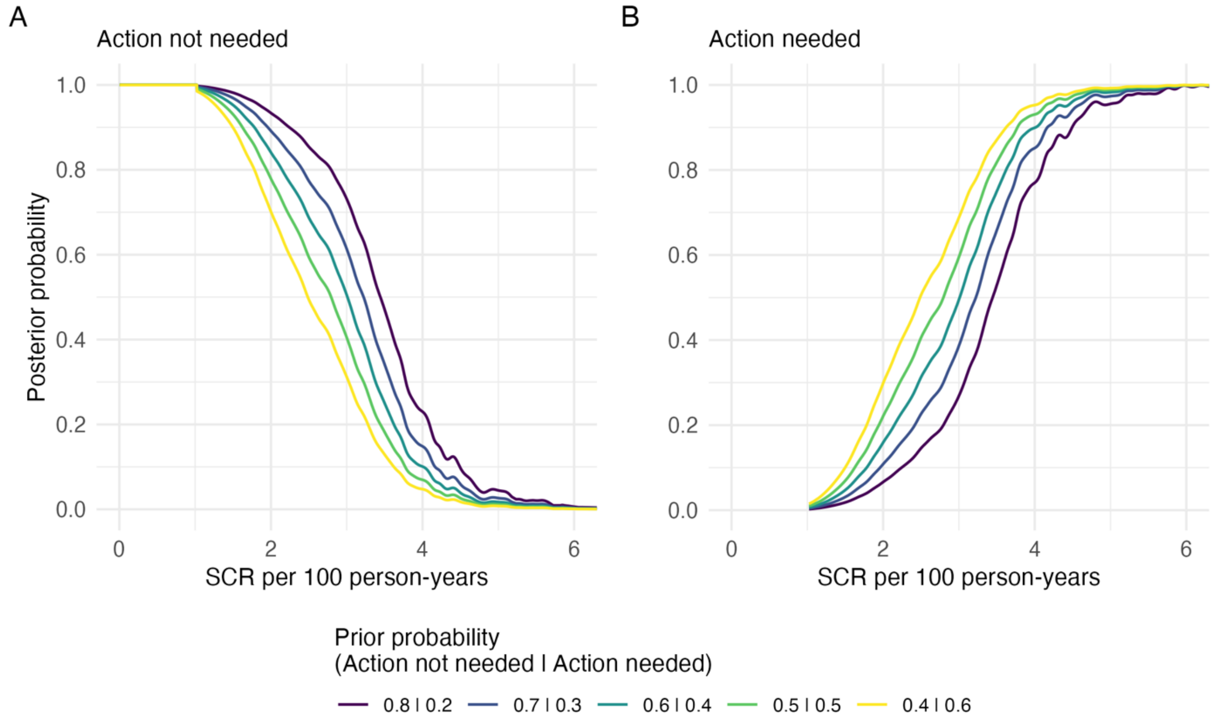

**Fig S5. Comparison of Pgp3 IgG seroprevalence and seroconversion rate (SCR) estimates using different age ranges.** Seroprevalence and SCR estimated at evaluation unit level in 43 surveys that used a population-based survey protocol for district or subdistrict level populations of 1–9-year-olds (n=14,620 aged 1–3 years, n=25,492 aged 1–5 years, n=43,606 aged 1–9 years). In all panels the dashed line indicates the 1:1 relationship and the solid line is a locally weighted regression fit. Blue points denote the 1–9-year-old vs 1–5-year-old comparison and red points mark the 1–3-year-old vs 1–5-year-old comparison. **A)** Seroprevalence estimated among 1–9-year-olds versus the same value estimated among 1–5-year-olds. **B)** SCR estimated among 1–9-year-olds versus the same value estimated among 1–5-year-olds. **C)** Seroprevalence estimated among 1–3-year-olds versus the same value estimated among 1–5-year-olds. **D)** SCR estimated among 1–3-year-olds versus the same value estimated among 1–5-year-olds. Although 1–5-year-old seroprevalence is systematically lower than seroprevalence among 1–9-year-olds (panel A) and higher than when estimated among 1–3-year-olds (panel C), SCR is broadly consistent when estimated using the different age ranges (panels B, D).

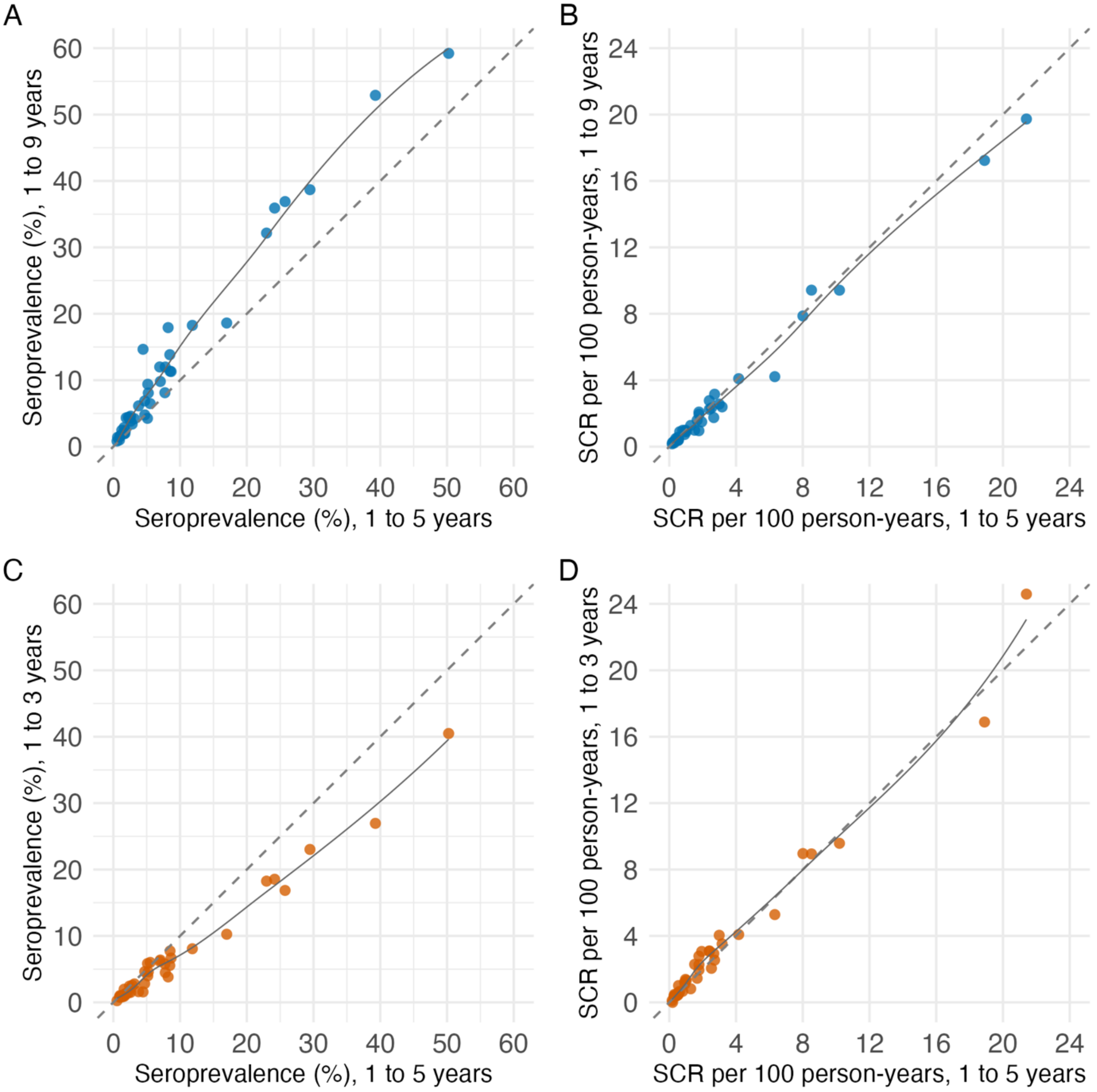

**Fig S6. Comparison of (Pgp3 + Ct694) IgG responses versus Pgp3 alone for seroprevalence and seroconversion rate (SCR) estimation.** Seroprevalence and SCR per 100 person-years (PY) estimated at the evaluation unit level in 36 surveys that had measured IgG responses to both Pgp3 and Ct694 antigens in 1–5-year-olds. In all panels the dashed line indicates the 1:1 relationship. The histograms summarize the differences between estimates with double versus single antigen.

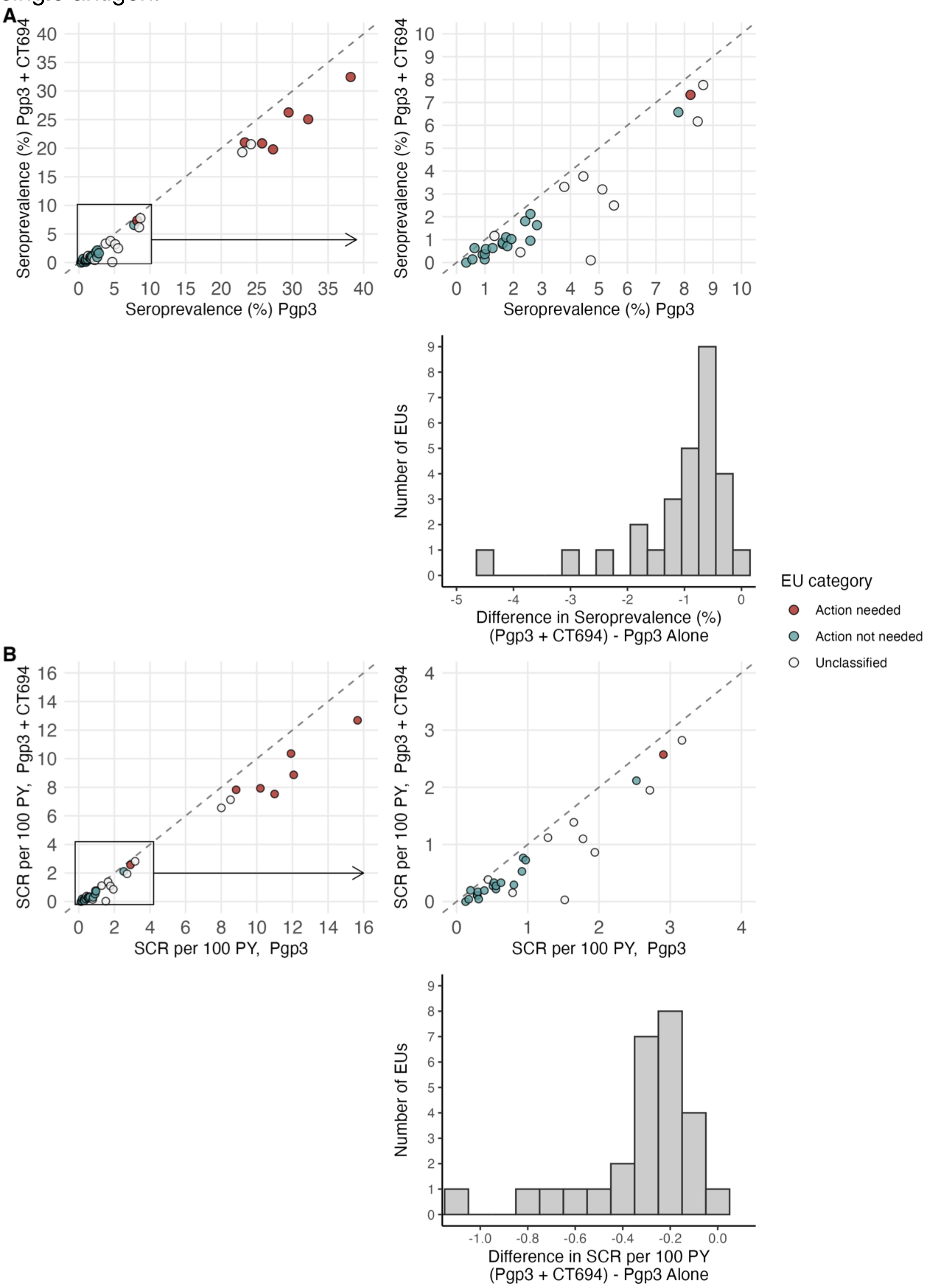

**Fig S7. Comparison of the seroconversion rate (SCR) estimated using a reversible catalytic model allowing for seroreversion with a model that assumes no seroreversion**. Allowing for seroreversion, here assumed to be 6 per 100 person-years based on previous estimates from longitudinal cohorts in near elimination transmission settings, scales the SCR linearly but does not change the rank order in estimates. Estimates are colored by evaluation unit (EU) trachoma programmatic decision category used in the main analyses (N=48 EUs). The dashed line indicates a 1:1 relationship and the solid line indicates a linear regression fit, with intercept 0 and slope 1.15 (adjusted $R^2$=1). The right panel provides a zoomed-in view of SCR estimates with values below 5 per 100 person-years.

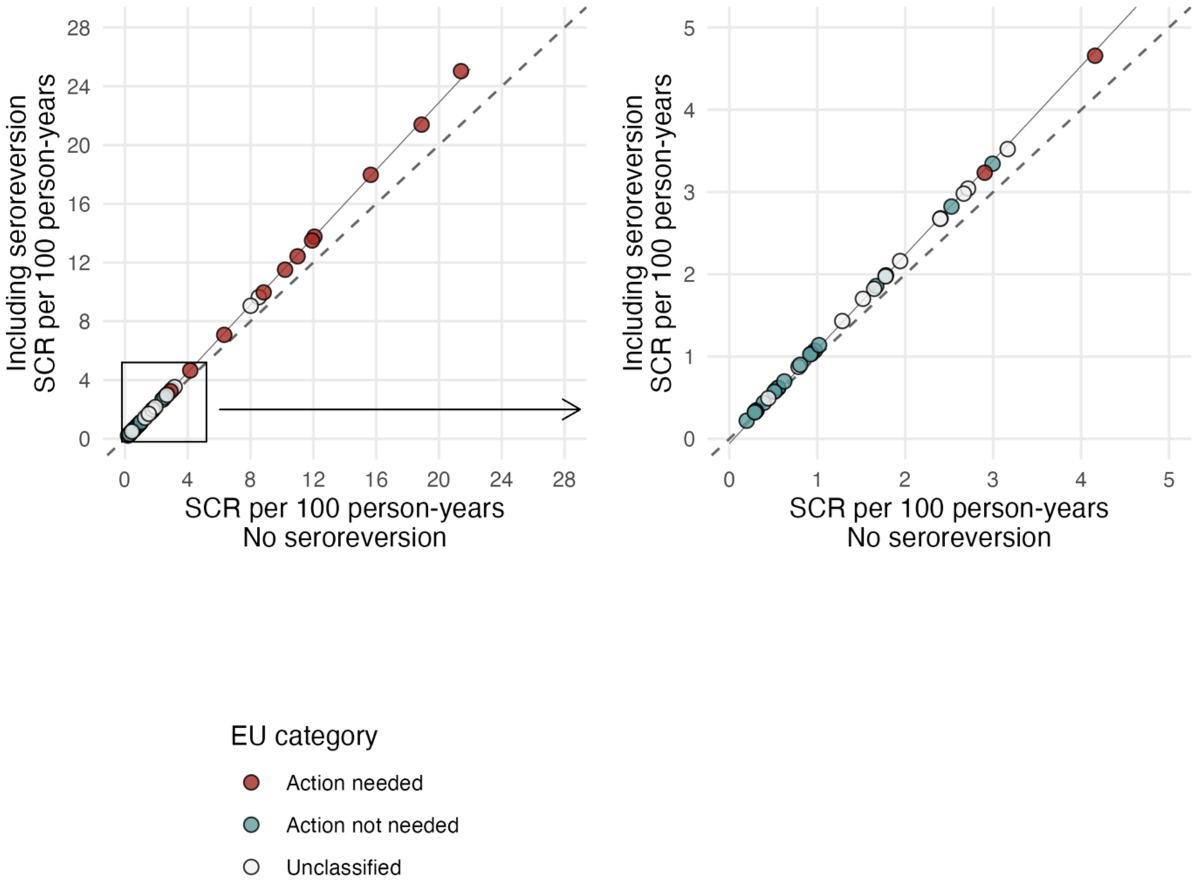

**Fig S8. Comparison of Bayesian Monte Carlo Markov Chain (MCMC) estimator versus maximum likelihood with generalized linear models (GLM) estimator.** Comparison of Pgp3 IgG seroprevalence (A) and seroconversion rate (B) estimated using MCMC and GLM estimators. Evaluation units (EU) are colored by transmission categories used in the main analysis (N=48 EUs). Dashed lines indicate the 1:1 relationship and right panels provide a zoomed view to estimates for populations at low levels of transmission.

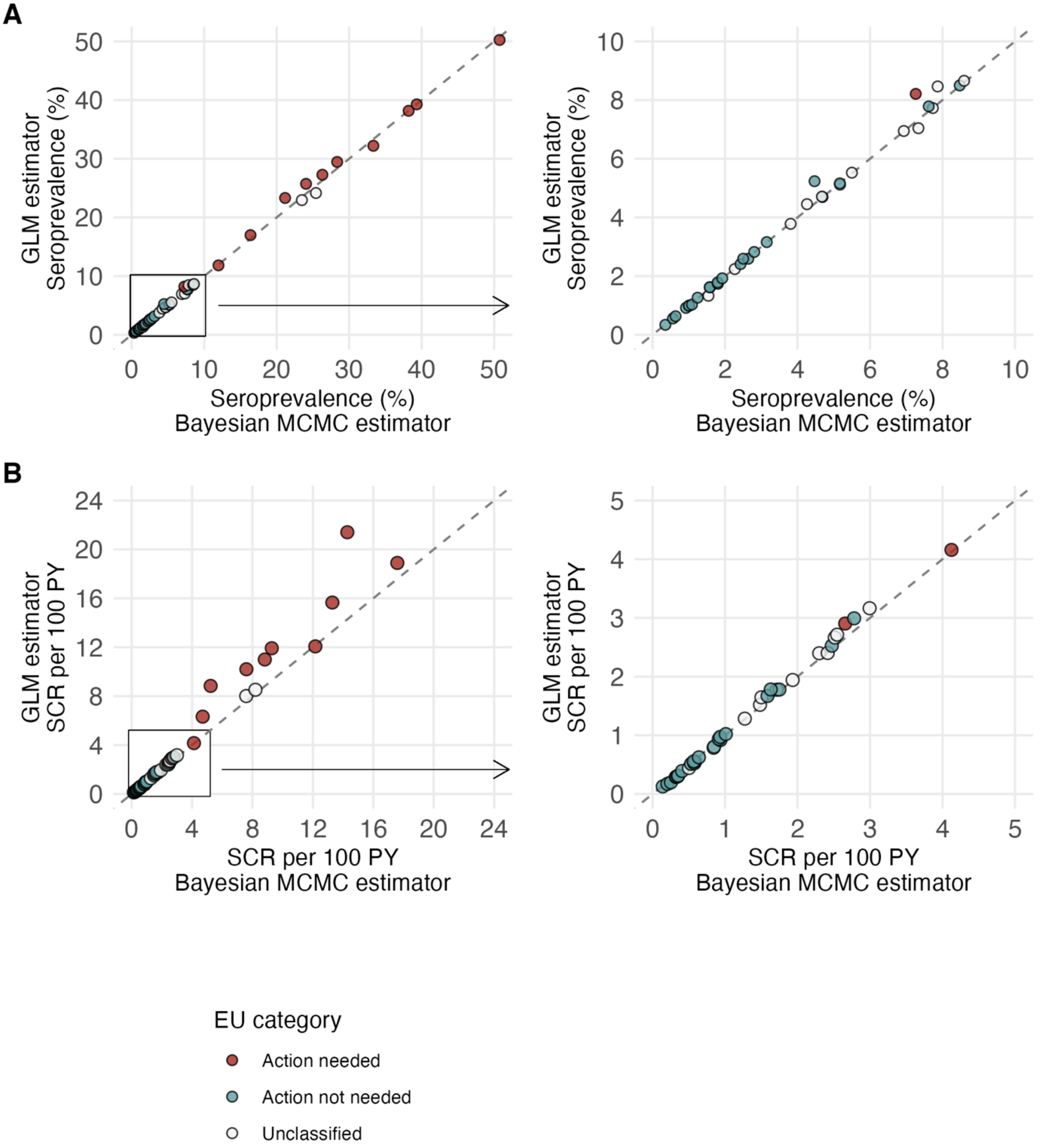

PY: person-years

**Fig S9. Comparison of seroprevalence estimates with and without age standardization.** Seroprevalence estimated at the evaluation unit level for three different age ranges in 39 evaluation unit surveys that used a population-based survey protocol for district or subdistrict level populations of 1–9-year-olds (n=14,620 aged 1–3 years, n=25,492 aged 1–5 years, n=43,606 aged 1–9 years). The results illustrate that age standardization (assuming uniform age distribution) has very little influence on seroprevalence estimates. **(A)** Estimates for all 43 evaluation units. The dashed line indicates the 1:1 relationship. The bounded box indicates the zoom window for panel in B. **(B)** Estimates limited to the 0–20% seroprevalence region to better visualize the data for populations at low levels of transmission.

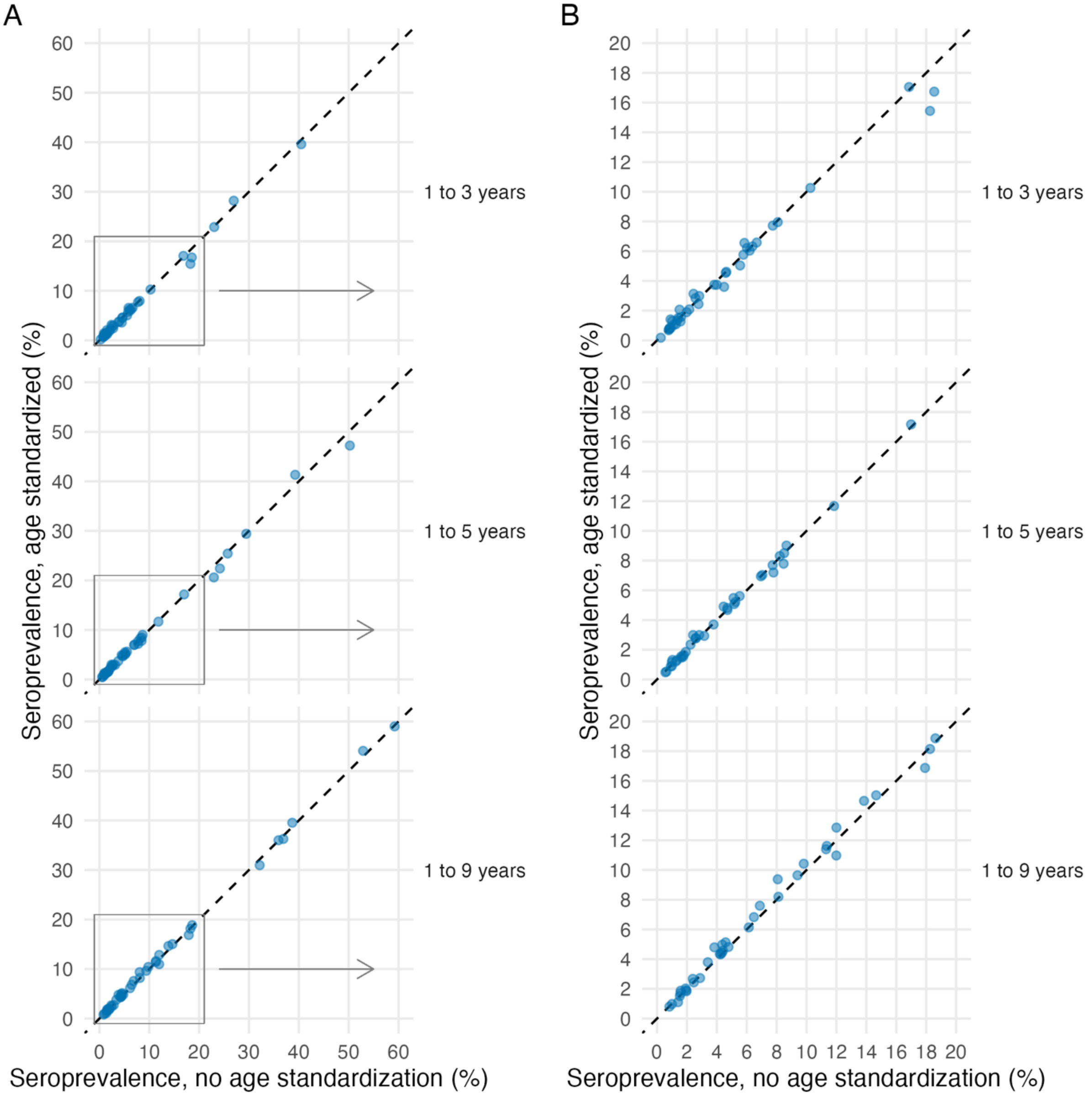
